# Simultaneous CBF and CMRGlu as imaging biomarkers for differential treatments in ICA/MCA steno-occlusive disease: a retrospective study

**DOI:** 10.64898/2026.02.05.26345710

**Authors:** Bixiao Cui, Yu Lu, Min Wang, Yi Shan, Jie Ma, Tao Wang, Yan Ma, Xiehui Jiang, Jie Lu

## Abstract

**BACKGROUND:** Steno-occlusive diseases of the internal carotid artery (ICA) or middle cerebral artery (MCA) can lead to hemodynamic impairment, yet conventional imaging often fails to reflect metabolic dysfunction. Integrated positron emission tomography and magnetic resonance imaging (PET/MRI) allows simultaneous assessment of cerebral blood flow (CBF) and glucose metabolism. This study compared baseline perfusion and metabolic characteristics between patients receiving medical therapy or extracranial–intracranial (EC-IC) bypass surgery.

**METHODS:** This retrospective study enrolled 34 patients with unilateral ICA/MCA stenosis or occlusion confirmed by digital subtraction angiography. All patients underwent ^18^F-FDG PET/MRI before treatment. Glucose metabolism was quantified using the cerebral metabolic rate of glucose (CMRGlu) from dynamic PET and the standard uptake value ratio (SUVR) from static PET. CBF was measured using three-dimensional arterial spin labeling with post-labeling delays of 2.0 and 2.5 seconds. Perfusion and metabolic parameters were compared across vascular territories.

**RESULTS:** Baseline clinical characteristics and long-term outcomes did not differ between groups (all *P*>0.05). Cerebral blood flow was similar across all arterial territories and post-labeling delays, with no hemispheric asymmetry detected (all *P*>0.05). In contrast, glucose metabolism was significantly lower in the surgical group, with reduced CMRGlu in the ischemic middle cerebral artery (23.58±7.46 vs 18.82±5.04μmol/100g^-1^/min^-1^, *P*=0.037) and anterior cerebral artery territories (26.37±8.76 vs 20.71±5.78μmol/100g^-1^/min^-1^, *P*=0.034). No differences were observed in the posterior cerebral artery or in SUVR across all regions (all *P*>0.05).

**CONCLUSIONS:** Despite similar perfusion profiles, the surgical group demonstrated lower glucose metabolism, suggesting that metabolic imaging may aid in identifying patients who could benefit from revascularization.

## Main

Ischemic stroke is a major global public health problem because of its high mortality and long-term disability.^1^ In China, the burden of stroke-related mortality is particularly pronounced, accounting for approximately one-third of global stroke-related deaths. Among the numerous causative factors contributing to ischemic stroke, stenosis and occlusion of the internal carotid artery (ICA) and middle cerebral artery (MCA) represent one of the primary etiologies.^2–4^ Patients with ICA or MCA stenosis or occlusion typically experience chronic or subacute hypoperfusion, which can induce neurological symptoms such as dizziness, paresthesia, and speech impairment. Persistent or worsening ischemia may eventually lead to irreversible brain injury and long-term disability. This severely compromises patient prognosis and quality of life while imposing heavy burdens on family caregiving and societal healthcare resources.^5, 6^ The optimal treatment strategy for this condition remains a subject of long-standing clinical debate, with no established consensus.^7, 8^ For patients with cerebral ischemia caused by symptomatic ICA or MCA steno-occlusive, two primary clinical approaches are currently employed: first, medical management centered on antiplatelet therapy, lipid-lowering agents, and comprehensive risk factor control; second, endovascular-to-intracranial (EC-IC) bypass surgery aimed at restoring cerebral blood flow. Although both approaches are widely used clinically, questions regarding the efficacy differences between the two treatment strategies and the precise selection of suitable patient populations remain unresolved, posing challenges for clinical decision making.

Existing clinical studies indicate no significant statistical difference between the two approaches in terms of the risk of stroke or death within 30 days, or the risk of ipsilateral ischemic stroke between 30 days and 2 years.^9^ However, in certain subgroups such as those with MCA occlusion or severe hypoperfusion, some studies suggest that bypass surgery may provide better prognostic improvement.^10^ This contradictory outcome reflects the current clinical lack of effective tools to precisely identify patients who truly benefit from reperfusion, leading to insufficient individualization in treatment decisions. The core issue of this clinical dilemma lies in the limitations of traditional assessment systems. Conventional imaging evaluations primarily rely on the degree of vascular stenosis or single perfusion metrics; however, perfusion levels alone may not fully reflect the compensatory capacity and functional state of brain tissue. Growing evidence indicates that during chronic or subacute hypoperfusion, brain tissue can maintain functional stability through mechanisms such as enhanced collateral circulation and cellular-level alterations in energy substrate utilization and metabolic pathways-a phenomenon termed “metabolic adaptation”. Therefore, even when blood flow perfusion levels are similar across patients, their metabolic states and functional reserves may exhibit significant variability. Some patients demonstrate preserved metabolism despite reduced blood flow, while others show markedly impaired glucose metabolism even without significant blood flow reduction, indicating a more fragile neural functional state.

The development of PET/MRI integrated imaging technology offers a potential solution to this challenge. This technology enables simultaneous acquisition of glucose metabolism information from ^18^F-fluorodeoxyglucose (FDG) PET and blood flow parameters from MRI perfusion sequences such as ASL during a single examination, yielding complementary data on cerebral hemodynamics and metabolic status in a single phase. Specifically, ¹⁸F-FDG PET can be used for quantitative or semi-quantitative characterization of cerebral glucose metabolism (e.g. CMRGlu or SUVR), reflecting neuronal energy utilization. Previous studies in patients with ICA or MCA steno-occlusive have observed that reduced glucose metabolism in certain regions may precede overt structural damage, suggesting FDG imaging exhibits high sensitivity in detecting functional abnormalities associated with chronic hypoperfusion.^11^ More importantly, PET/MRI studies suggest that cerebral blood flow (CBF) and cerebral glucose metabolism rate (CMRGlu) do not always change synchronously under chronic ischemic conditions. The presence of “perfusion-metabolism mismatch” in certain regions may directly reflect variations in brain tissue compensatory capacity.^12^

Therefore, this study employs a retrospective observational design to analyze patients with internal carotid artery/middle cerebral artery stenosis or occlusion who underwent either medical therapy or EC-IC bypass surgery. The objectives are: (1) to compare cerebral blood flow and glucose metabolism characteristics at baseline admission between the two groups, determining whether differences exist in baseline perfusion and metabolic status between patients receiving different treatments; (2) To explore the association between these baseline imaging characteristics and patient treatment selection as well as potential prognosis.

## METHODS

### Study Population

This study employed a retrospective, observational single-center cohort design, enrolling patients with unilateral ICA or MCA stenotic or occlusive disease who received standardized long-term management at our institution between 2017 and 2022. Following clinical evaluation, the final treatment modality was determined by clinicians based on the patient’s condition and relevant examination results, and jointly decided by the patient and family after thorough communication. Based on the final treatment choice, patients were assigned to either the surgical group (extracranial-intracranial bypass surgery combined with medical therapy) or the medical group (medical therapy alone). Inclusion criteria include: (1) Digital subtraction angiography (DSA) confirmed unilateral ICA or MCA steno-occlusive; (2) Transient ischemic attack (TIA) or ischemic stroke occurring within the past 12 months in the territory supplied by the affected artery, with the most recent stroke episode ≥3 weeks prior to imaging and stable neurological function for ≥1 month before treatment; (3) Completion of PET/MRI imaging prior to treatment; (4) Standardized clinical assessment by a neurologist at admission, recording baseline scores on the NIH Stroke Scale (NIHSS) and modified Rankin Scale (mRS); (5) All patients completed at least 2 years of follow-up (median 1857 days, range: 801–2685 days). Exclusion criteria included: (1) loss to follow-up or inability to obtain complete clinical follow-up data; (2) refusal or inability to complete PET/MRI imaging; (3) poor PET/MRI image quality due to motion artifacts, metal artifacts, or reconstruction errors, rendering reliable quantitative analysis impossible; (4) Concurrent central nervous system diseases (e.g., neurodegenerative disorders, brain tumors, epilepsy) likely to significantly affect cerebral metabolism or blood flow. This study protocol was approved by the Ethics Committee of Xuanwu Hospital (Ethics No. [2025]286-002). The patient screening process is illustrated in Figure 1. Between 2017 and 2022, 57 patients meeting the diagnostic criteria for unilateral ICA or MCA steno-occlusive were screened. Based on inclusion and exclusion criteria, 23 cases were excluded (11 lost to follow-up, 7 unable to complete or refused PET/MRI examination, 5 with poor image quality). Ultimately, 34 patients were included in the analysis: 17 in the surgical group and 17 in the medical group.

**Figure 1.**
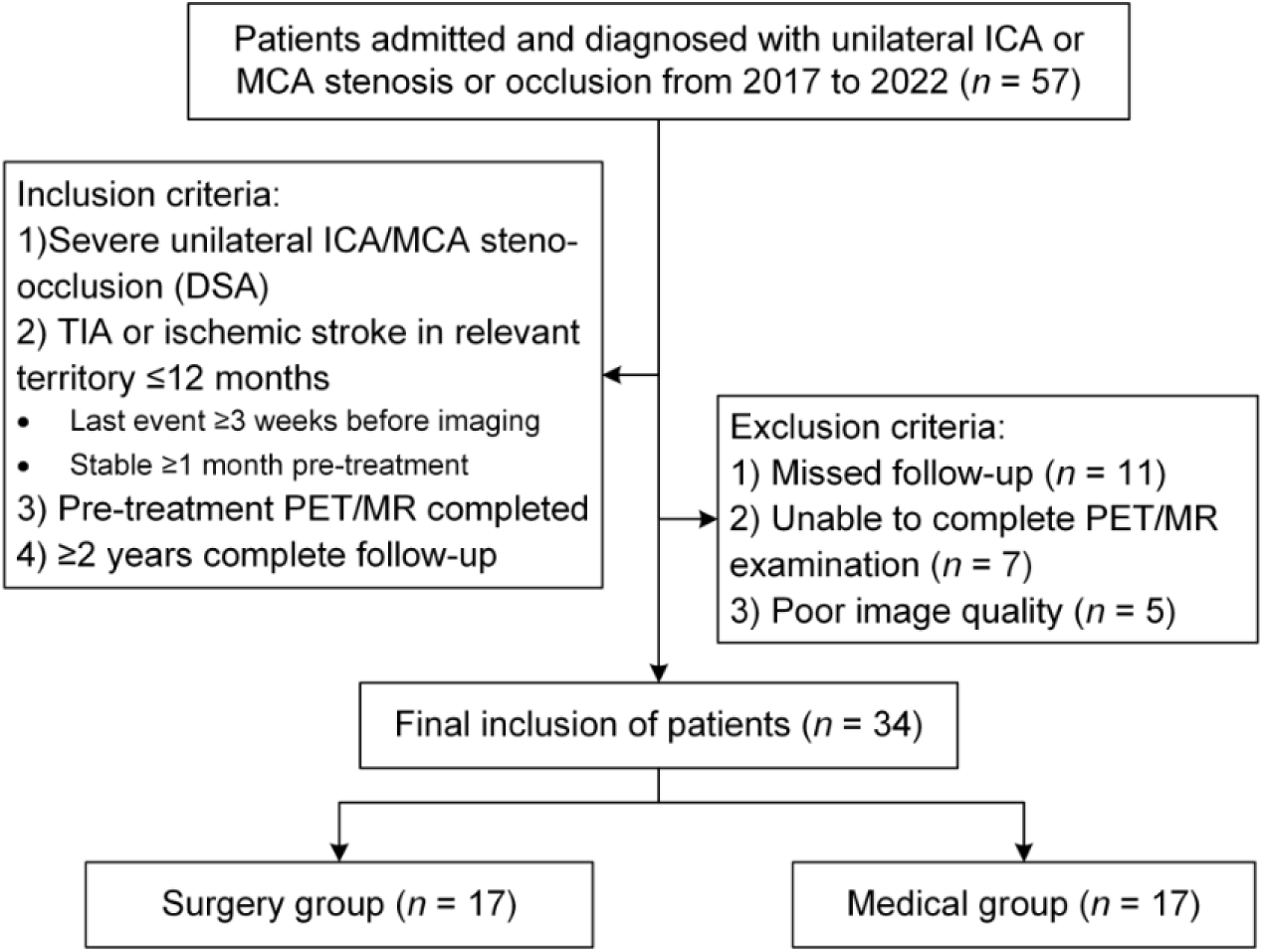
Patient Inclusion and Exclusion Flowchart. Abbreviations: ICA, Internal carotid artery; MCA, Middle cerebral artery; PET/MRI, Positron emission tomography and magnetic resonance imaging.

### PET/MRI Scanning Protocol

All subjects underwent simultaneous PET and MRI examinations using an integrated TOF-PET/MRI system (Signa, GE Healthcare). All subjects fasted for at least 6 hours prior to the examination, and blood glucose levels were measured for each subject to ensure serum glucose concentrations remained below 8 mmol/L. Scans were performed at rest without any task instructions. A 19-channel head neck combined coil was used to achieve high signal to noise ratio for PET/MRI imaging. Subjects were positioned supine, centered within the field of view, and instructed to maintain a calm state with eyes closed. PET and MRI data were acquired synchronously.

Each subject received an intravenous administration of [^18^F] FDG at a dose of 3.7 MBq/kg. PET/MRI acquisition commenced approximately 50 minutes after tracer injection, and PET emission data were collected for 10 minutes. PET images were corrected for attenuation using MRI-based Dixon sequences provided by the manufacturer. Image reconstruction was performed using a time-of-flight point spread function ordered subset expectation maximization (TOF-PSF-OSEM) algorithm with 8 iterations and 32 subsets, followed by application of a 3mm Gaussian post-reconstruction filter. The reconstructed images had a matrix size of 192 × 192, a field of view of 35 × 35 cm^2^, and a voxel resolution of 1.82 × 1.82 × 2.78 mm^3^.

PET and MRI datasets were obtained simultaneously. The MRI scanning sequences included 3D T1, MRA, T2 flair, and 3D ASL (PLD=1.5s, PLD=2.0s). Specific scanning parameters and protocols are detailed in Table S1 of the supplementary materials.

### Image Processing

MRI images were evaluated by two radiology attending physicians with 5-10 years of experience, who were required to report any structural abnormalities in subcortical white matter and gray matter or alterations in T2WI signal intensity. Disagreements were resolved by a third radiology chief physician. For patients with negative MRI findings or suspected minor abnormalities, MRI images were re-evaluated under ^18^F-FDG PET/MRI guidance. Infarct areas were individually annotated by two experienced radiologists from the Department of Radiology and Nuclear Medicine based on T2 flair images, then standardized to Montreal Neurological Institute (MNI) space. Cerebral glucose metabolic rate (CMRGlu) parameter images (μmol/100g^-1^/min^-1^) were extracted from dynamic FDG PET using a fully automated workflow, as previously described in the literature.^13, 14^ These were then normalized to MNI space and smoothed using a 6 mm full width at half maximum (FWHM) isotropic Gaussian kernel to reduce noise. Static FDG PET images were first registered to T1-weighted MRI, normalized to MNI space, and similarly smoothed with a 6 mm FWHM Gaussian kernel. SUVR were calculated using the cerebellopontine region as the reference area. CBF images were automatically generated from ASL images using the standard ASL quantification model previously described.^15^ These images underwent registration with T1-weighted MRI, normalization to MNI space, and 6 mm FWHM Gaussian kernel smoothing for subsequent analysis. After removing infarct regions from all processed images, they were matched to cerebral arterial supply territories. CBF, CMRGlu, and SUVR values were calculated for the affected and contralateral territories of the ACA, MCA, and posterior cerebral artery (PCA), respectively.

### Statistical Analysis

All statistical analyses and graphing in this study were performed using SPSS 26.0 and GraphPad Prism 10.0. Normality of continuous variables was assessed using the Shapiro-Wilk test. Normally distributed variables are expressed as mean ± standard deviation, with intergroup comparisons performed using independent samples t-tests; non-normally distributed variables are expressed as median (IQR), with intergroup comparisons performed using the Mann-Whitney *U* test. Categorical variables were analyzed using the *χ*^2^ test or Fisher’s exact test. Imaging parameters (CBF, CMRGlu, SUVR) were compared between the surgical and medical groups separately for each supply region (ACA, MCA, PCA). Comparisons between the affected and unaffected sides within each group were performed using paired *t*-tests or Wilcoxon signed-rank tests based on data distribution characteristics. All statistical tests were two-sided, and *P* < 0.05 was considered statistically significant.

## RESULTS

### Baseline Clinical Characteristics

From May 2017 to August 2022, a total of 34 patients were diagnosed with unilateral internal carotid artery or middle cerebral artery stenotic occlusive disease via DSA. Among them, 17 (50.00%) underwent temporal superficial artery-middle cerebral artery bypass surgery, while 17 received medical therapy. The two groups were generally comparable in baseline demographic and clinical characteristics (Table 1). No significant differences were observed between the medical and surgical groups in age (47.06 ± 10.32 years versus 49.88 ± 7.24 years, *P*=0.362), gender distribution (41.2% female versus 17.6%, *P*=0.132), height, weight, or body mass index (BMI) (all *P*>0.05). Differences in lesion side (left/right: 6/11 versus 8/9, *P*=0.486) and affected artery distribution (ICA/MCA: 4/13 versus 8/9, *P*=0.151) were also not statistically significant between the two groups.

**Table 1.**
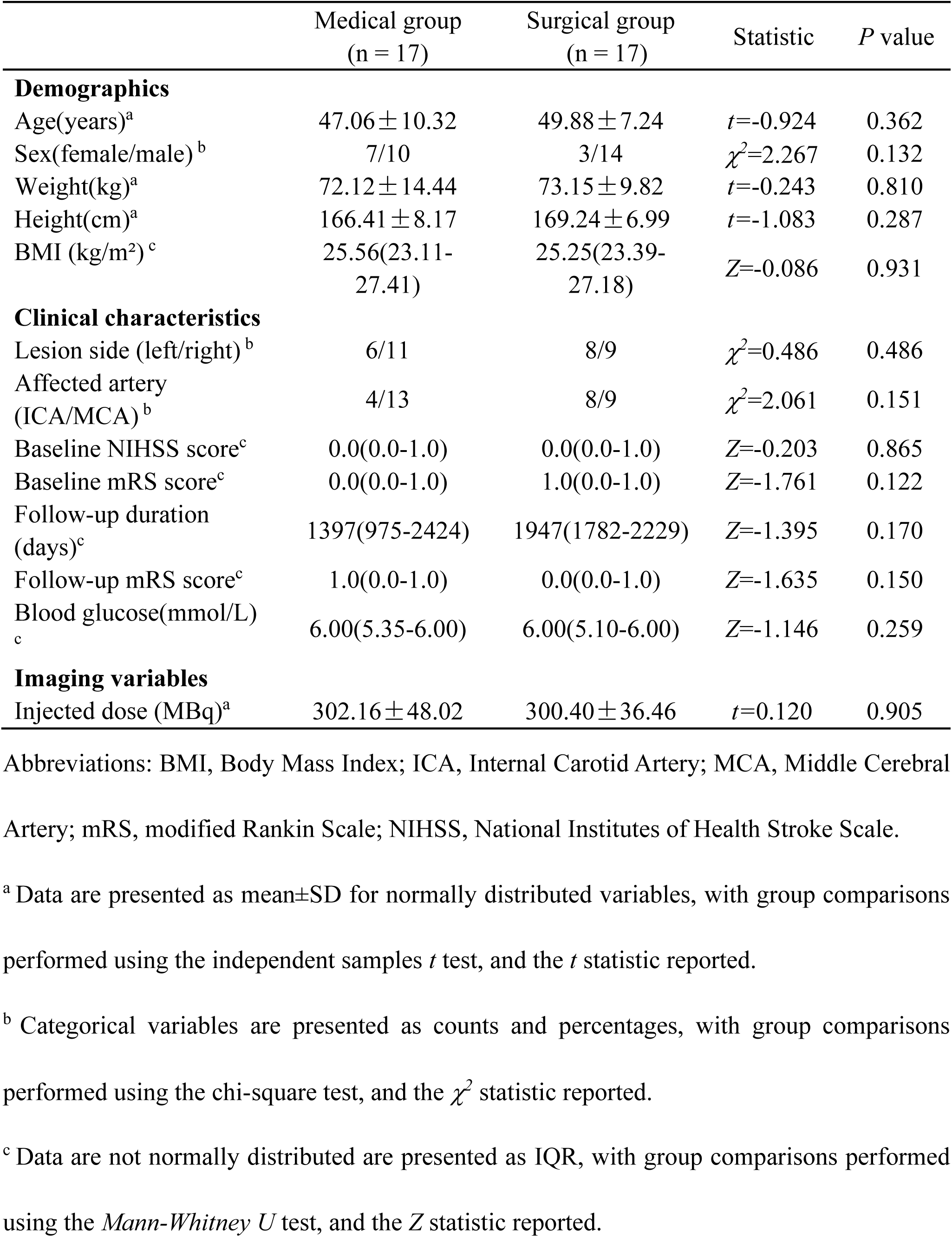
Baseline Demographics and Clinical Characteristics of the Patients

|  | Medical group<br>(n = 17) | Surgical group<br>(n = 17) | Statistic | P value |
| --- | --- | --- | --- | --- |
| **Demographics** |  |  |  |  |
| Age(years)<sup>a</sup> | 47.06 ± 10.32 | 49.88 ± 7.24 | $t=-0.924$ | 0.362 |
| Sex(female/male)<sup>b</sup> | 7/10 | 3/14 | $\chi^2=2.267$ | 0.132 |
| Weight(kg)<sup>a</sup> | 72.12 ± 14.44 | 73.15 ± 9.82 | $t=-0.243$ | 0.810 |
| Height(cm)<sup>a</sup> | 166.41 ± 8.17 | 169.24 ± 6.99 | $t=-1.083$ | 0.287 |
| BMI (kg/m<sup>2</sup>)<sup>c</sup> | 25.56(23.11-27.41) | 25.25(23.39-27.18) | $Z=-0.086$ | 0.931 |
| **Clinical characteristics** |  |  |  |  |
| Lesion side (left/right)<sup>b</sup> | 6/11 | 8/9 | $\chi^2=0.486$ | 0.486 |
| Affected artery<br>(ICA/MCA)<sup>b</sup> | 4/13 | 8/9 | $\chi^2=2.061$ | 0.151 |
| Baseline NIHSS score<sup>c</sup> | 0.0(0.0-1.0) | 0.0(0.0-1.0) | $Z=-0.203$ | 0.865 |
| Baseline mRS score<sup>c</sup> | 0.0(0.0-1.0) | 1.0(0.0-1.0) | $Z=-1.761$ | 0.122 |
| Follow-up duration<br>(days)<sup>c</sup> | 1397(975-2424) | 1947(1782-2229) | $Z=-1.395$ | 0.170 |
| Follow-up mRS score<sup>c</sup> | 1.0(0.0-1.0) | 0.0(0.0-1.0) | $Z=-1.635$ | 0.150 |
| Blood glucose(mmol/L)<br><sup>c</sup> | 6.00(5.35-6.00) | 6.00(5.10-6.00) | $Z=-1.146$ | 0.259 |
| **Imaging variables** |  |  |  |  |
| Injected dose (MBq)<sup>a</sup> | 302.16 ± 48.02 | 300.40 ± 36.46 | $t=0.120$ | 0.905 |

Abbreviations: BMI, Body Mass Index; ICA, Internal Carotid Artery; MCA, Middle Cerebral Artery; mRS, modified Rankin Scale; NIHSS, National Institutes of Health Stroke Scale.
<sup>a</sup> Data are presented as mean±SD for normally distributed variables, with group comparisons performed using the independent samples $t$ test, and the $t$ statistic reported.
<sup>b</sup> Categorical variables are presented as counts and percentages, with group comparisons performed using the chi-square test, and the $\chi^2$ statistic reported.
<sup>c</sup> Data are not normally distributed are presented as IQR, with group comparisons performed using the *Mann-Whitney U* test, and the $Z$ statistic reported.

Baseline neurological function scores were comparable between the two patient groups, including baseline NIHSS (0.0[0.0–1.0] versus 0.0[0.0–1.0], *P*=0.865) and baseline mRS (0.0[0.0–1.0] versus 1.0[0.0–1.0], *P*=0.122). There was also no significant difference in follow-up duration between the two groups (1397[975–2424] vs.1947[1782–2229], *P*=0.170). Comparison of mRS scores at follow-up also showed no difference (1.0[0.0–1.0] versus 0.0[0.0–1.0], *P*=0.150). Further analysis of functional changes from baseline to follow-up revealed a slight increase in mRS in the medication group and a mild decrease in the surgery group. However, neither group demonstrated significant functional deterioration or improvement (both *P*> 0.05). Changes in mRS from baseline to follow-up for both groups are shown in Figures 2A and 2B.

**Figure 2.**
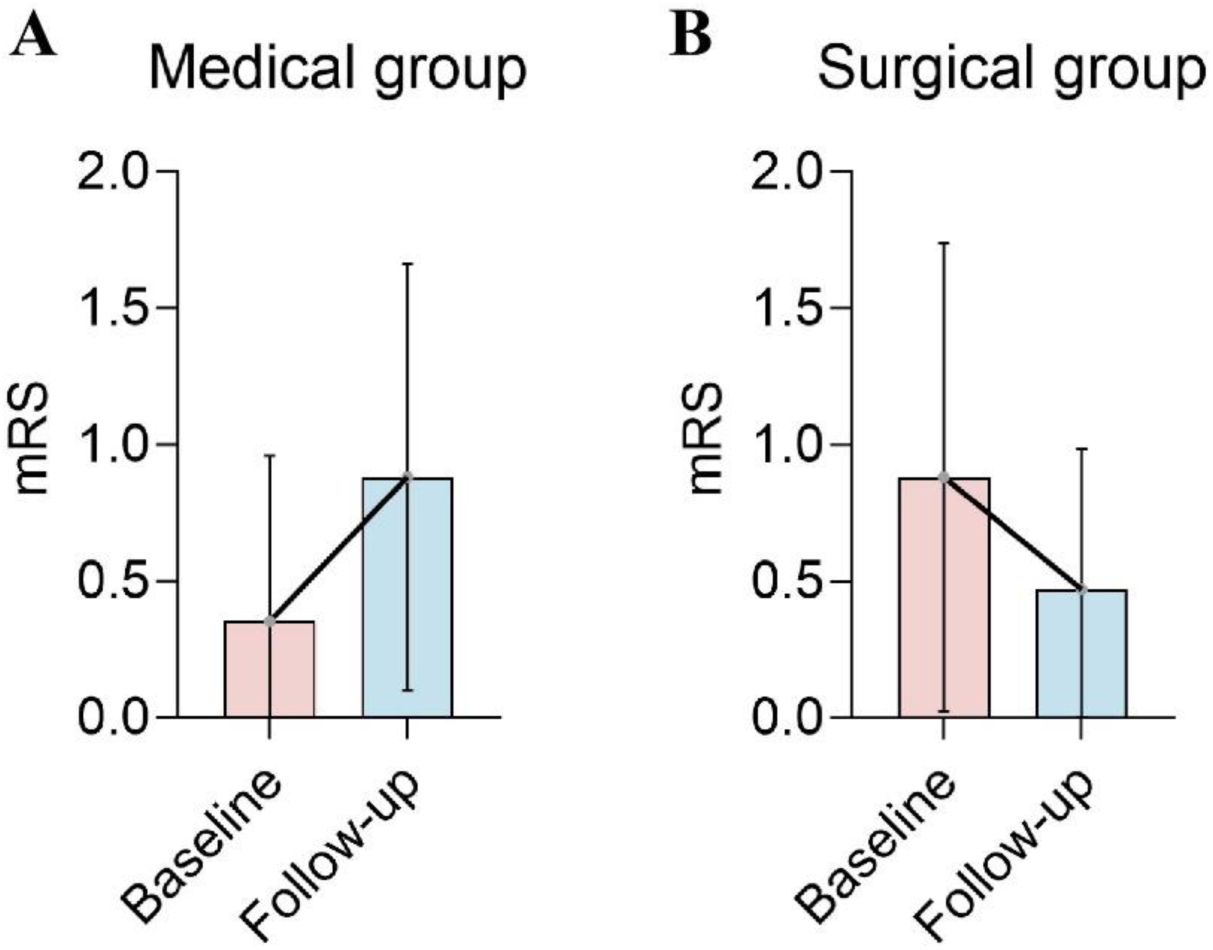
mRS scores in patients receiving medical or surgical treatment. (A) Changes in mRS scores from baseline to follow-up in the medical group. (B) Changes in mRS scores from baseline to follow-up in the surgical group. Abbreviations: mRS, Baseline and follow-up modified Rankin Scale.

Additionally, the two groups showed no significant differences in blood glucose levels (6.00[5.35–6.00] mmol/L versus 6.00[5.10–6.00] mmol/L, *P*=0.259) and injection dose (302.16±48.02 MBq versus 300.40±36.46 MBq, *P*=0.905). Representative multimodal PET/MRI images from one patient in the surgical group and one patient in the medical group are provided in Figure S1 and Figure S2 in the Supplementary Materials.

### Blood Flow Analysis

Blood flow analysis results between the two groups showed that in the ischemic side, no significant differences in CBF were observed between the medical and surgical groups in the major arterial supply regions (MCA, ACA, PCA) under either PLD (2.0 s or 2.5 s) (all *P* > 0.05) (Table 2, Figures 3A and 3B). At PLD=2.0 s, CBF in the MCA, ACA, and PCA regions showed no significant differences between groups. At PLD=2.5 s, CBF in all regions of interests (ROI) also exhibited no statistically significant differences. Overall, the distribution trends of CBF on the ischemic side were similar between groups under both PLD conditions.

**Table 2.**
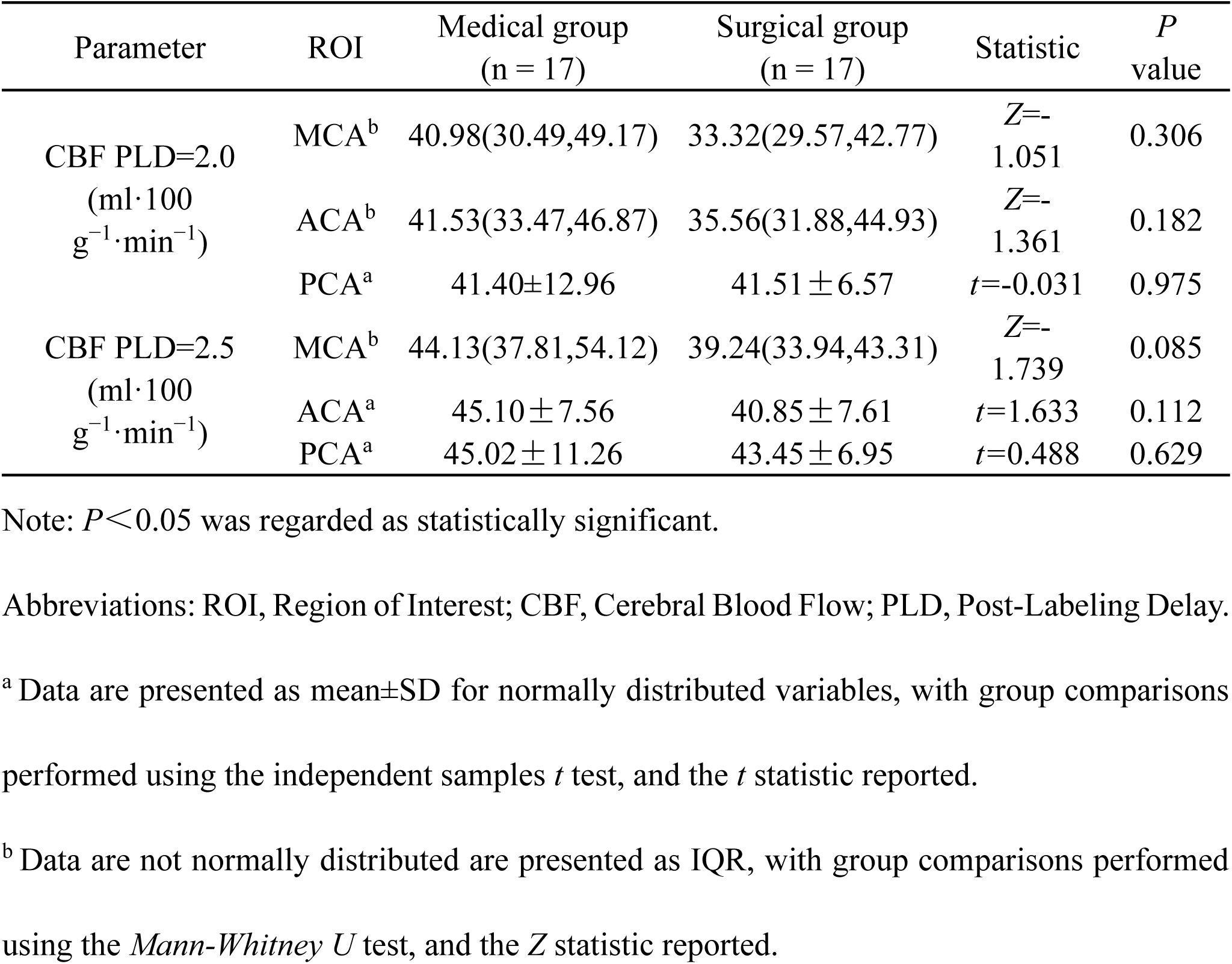
Comparison of Cerebral Blood Flow Between the Two Groups on the Ischemic Side

**Figure 3.**
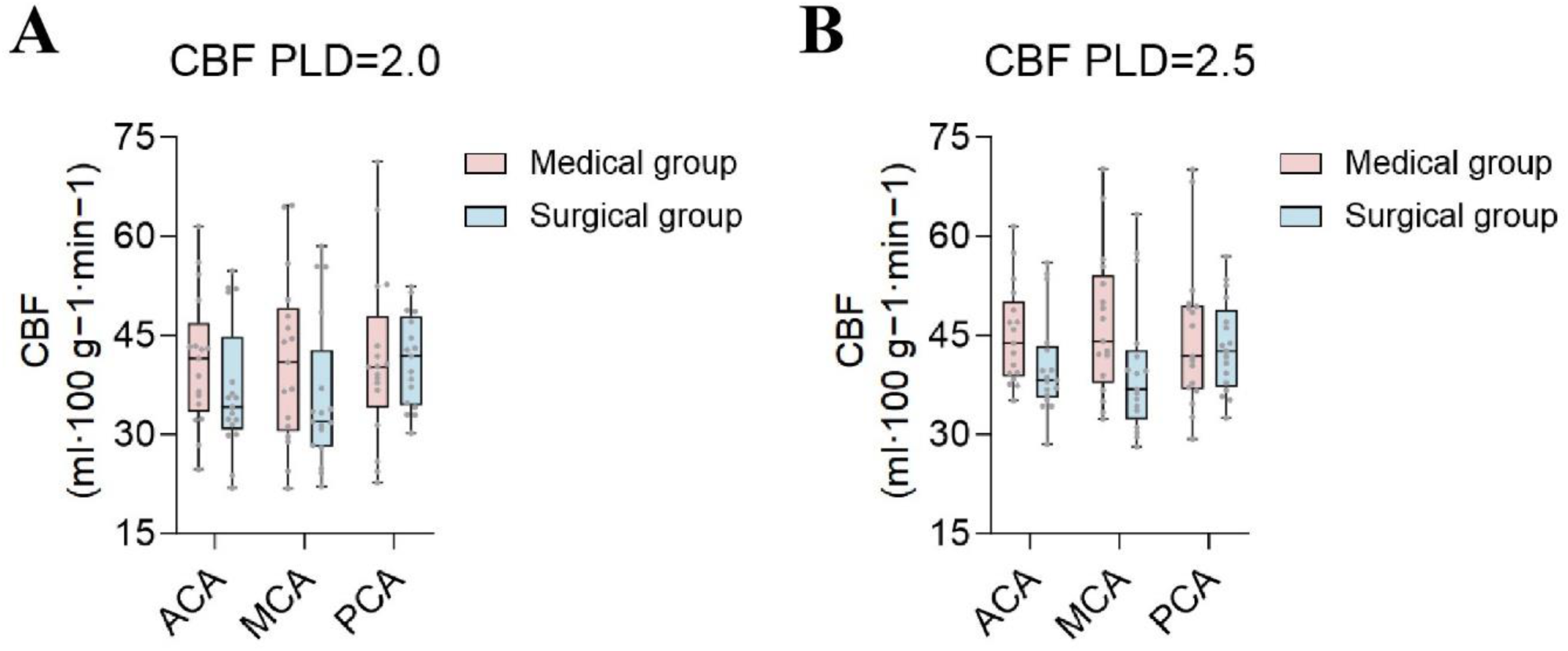
CBF comparisons between the medical and surgical groups across arterial territories on the ischemic side. (A) PLD=2.0 s. (B) PLD=2.5 s. Abbreviations: MCA, middle cerebral artery; ACA, anterior cerebral artery; PCA, posterior cerebral artery.

To further evaluate blood flow in the contralateral side across different groups, this study provides comparative CBF results for the contralateral side in both groups in the appendix (Table S2). The results indicate that for both PLD=2.0 s and 2.5 s, the differences in CBF across all ROIs in the contralateral side were not significant (all *P* > 0.05).

Additionally, this study compared CBF between the infarct side and the contralateral side in both the medical group and the surgical group (Table S3, Table S4). In the medical group, no significant differences in CBF were observed between the infarct side and the contralateral side for any ROI under either PLD condition (all *P* > 0.05). In the surgical group, no statistically significant differences in CBF were observed between the infarct side and the contralateral side (all *P* > 0.05).

In summary, consistent trends in CBF were observed across both the ischemic and non-ischemic sides, as well as in both the medical and surgical groups, within the MCA, ACA, and PCA regions, with no significant differences detected. However, on the ischemic side, the surgical group exhibited overall lower CBF compared to the medical group. Specifically, CBF in the MCA region decreased by approximately 19% and 11% at PLD=2.0 s and 2.5 s, respectively; The ACA region showed reductions of approximately 14% and 9%, respectively; the PCA region exhibited a mild decrease (approximately 0–3%).

### Metabolic Analysis

Metabolic analysis results between the two groups revealed intergroup differences in CMRGlu across distinct arterial supply regions on the ischemic side. Compared to the surgery group, the drug group exhibited significantly higher CMRGlu in the MCA region (*P* = 0.037), with similar findings observed in the ACA region (*P* = 0.034) (Table 3, Figure 4A). No significant intergroup differences were observed in the PCA region (*P* > 0.05). No statistically significant differences in SUVR were detected between the two groups in the MCA, ACA, or PCA regions (all *P* > 0.05) (Figure 4B).

**Table 3.**
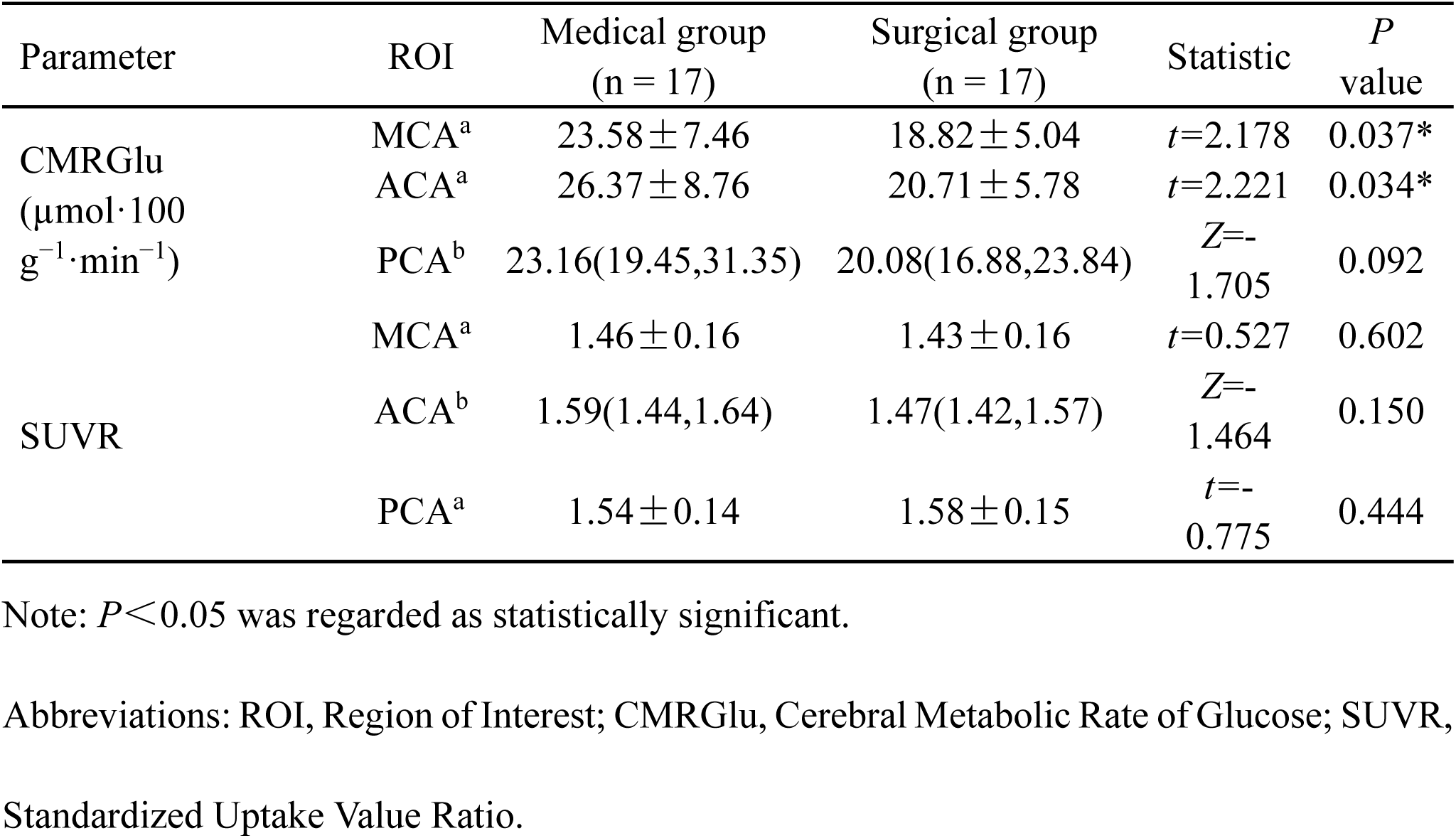
Comparison of Metabolic Parameters Between the Two Groups on the Ischemic Side

| Parameter | ROI | Medical group<br>(n = 17) | Surgical group<br>(n = 17) | Statistic | *P*<br>value |
| --- | --- | --- | --- | --- | --- |
| CMRGlu<br>(μmol·100<br>g<sup>-1</sup>·min<sup>-1</sup>) | MCA<sup>a</sup> | 23.58±7.46 | 18.82±5.04 | *t*=2.178 | 0.037* |
|  | ACA<sup>a</sup> | 26.37±8.76 | 20.71±5.78 | *t*=2.221 | 0.034* |
|  | PCA<sup>b</sup> | 23.16(19.45,31.35) | 20.08(16.88,23.84) | *Z*=-<br>1.705 | 0.092 |
| SUVR | MCA<sup>a</sup> | 1.46±0.16 | 1.43±0.16 | *t*=0.527 | 0.602 |
|  | ACA<sup>b</sup> | 1.59(1.44,1.64) | 1.47(1.42,1.57) | *Z*=-<br>1.464 | 0.150 |
|  | PCA<sup>a</sup> | 1.54±0.14 | 1.58±0.15 | *t*=-<br>0.775 | 0.444 |

Note: *P*<0.05 was regarded as statistically significant.
Abbreviations: ROI, Region of Interest; CMRGlu, Cerebral Metabolic Rate of Glucose; SUVR, Standardized Uptake Value Ratio.
<sup>a</sup> Data are presented as mean ± SD for normally distributed variables, with group comparisons performed using the independent samples *t* test, and the *t* statistic reported.
<sup>b</sup> Data are not normally distributed are presented as IQR, with group comparisons performed using the *Mann-Whitney U* test, and the *Z* statistic reported.

**Figure 4.**
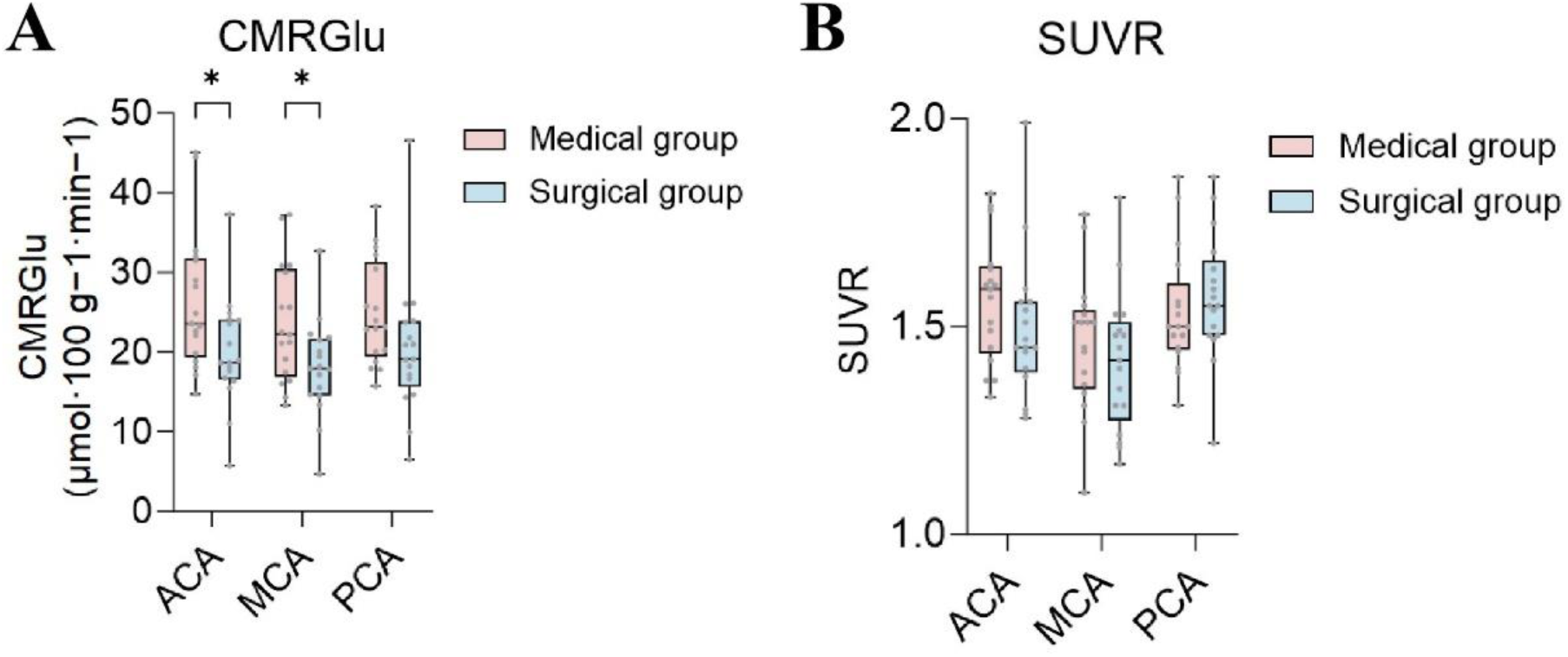
Comparison of metabolic parameters between the medical and surgical groups in different arterial territories on the ischemic side. (A) CMRGlu. (B) SUVR. \**P*<0.05.

On the unaffected side, no significant differences in CMRGlu or SUVR were observed between the two groups across all ROIs (Table S5, all *P* > 0.05). In internal comparisons between the surgical and medical groups, no significant differences in CMRGlu or SUVR were found between the infarct side and unaffected side in the MCA, ACA, and PCA regions (Table S6, Table S7, all *P* > 0.05).

In summary, metabolic indicators revealed: (1) Ischemic side: Glucose metabolism in the MCA and ACA was higher in the drug group than in the surgery group; no significant intergroup differences were observed in SUVR. (2) Contralateral side: Metabolic parameters were similar between groups. (3) Intragroup comparisons: Neither CMRGlu nor SUVR showed significant differences between the infarct and contralateral sides in either the drug or surgery group.

## DISCUSSION

This retrospective study compared baseline PET/MRI functional imaging characteristics between patients with chronic ICA/MCA stenosis-occlusion who underwent extracranial-intracranial (EC-IC) bypass surgery combined with medical therapy versus those receiving medical therapy alone. Patients underwent at least two years of clinical follow-up to assess long-term neurological outcomes. Results showed no significant difference in long-term neurological outcomes between the two groups. However, at the metabolic level, the surgical group exhibited significantly lower CMRGlu in the MCA and ACA supply areas compared to the medical group, while no statistically significant differences in CBF or SUVR were observed between the two groups. Furthermore, no differences were noted in any parameters within the unaffected PCA supply area.

The blood flow analysis in this study revealed that, regardless of whether PLD = 2.0 s or 2.5 s was used, the intergroup comparisons of CBF in both the ischemic and contralateral hemispheres between the surgical and medical groups did not reach statistical significance. However, it is noteworthy that in multiple ROIs on the normal side, the surgical group exhibited a trend toward lower CBF compared to the medical group, particularly consistent in the MCA distribution area. This non-significant yet consistent phenomenon may suggest weaker overall cerebral hemodynamic reserve in the surgical group. Previous studies have indicated that in intracranial atherosclerosis or chronic occlusive disease, some patients exhibit a marked decline in cerebrovascular reserve (CVR) even when CBF remains near normal. This suggests that CVR more sensitively reflects hemodynamic limitation than single CBF measurements, and that reduced blood flow reserve is closely associated with increased risks of ischemic events and stroke recurrence during follow-up.^15, 16^ Wang et al. findings similarly indicate that despite near-normal CBF in some chronic large vessel occlusion patients, prolonged mean transit time (MTT) and reduced relative cerebral blood flow (rCBF) significantly predict subsequent ischemic recurrence, suggesting static CBF alone cannot fully reveal underlying hemodynamic limitations.^17^ Further analysis of side-to-side differences revealed comparable CBF values between ischemic and non-ischemic sides in the medical group, whereas the ischemic side consistently showed lower CBF than the non-ischemic side in the surgical group. Although the differences were not statistically significant, this contrasting pattern of intra-group variation may reflect disparities in collateral circulation compensation capacity between the two groups: the medical group may have maintained ischemic-side CBF due to more robust collateral perfusion, whereas the surgical group likely exhibited more pronounced perfusion deficits or limited compensation, resulting in greater inter-hemispheric CBF disparities. Multiple studies indicate that in patients with chronic large vessel stenosis or occlusion, collateral circulation can partially sustain perfusion, allowing ischemic-side CBF to approach normal levels. However, this compensation does not reflect the true tissue state.^18, 19^ When collateral compensation is inadequate, patients are more prone to reduced perfusion reserve, delayed perfusion, and further functional decompensation, constituting a critical subgroup of potential beneficiaries for revascularization therapy.^7^

ASL technology itself harbors inherent limitations. When evaluating perfusion abnormalities in chronic occlusive diseases, ASL is significantly influenced by delayed arterial transit time (ATT). If labeled blood flow has not yet reached the capillary bed within the selected PLD, CBF may be underestimated or “pseudo normalized,” thereby masking true perfusion differences.^15^ Previous systematic reviews focusing on ASL further indicate that single-phase ASL accuracy markedly declines in patients with arterial stenosis or significant collateral circulation, whereas the temporal information from multi-PLD sequences more reliably depicts perfusion delays and true CBF status.^18^ The absence of significant intergroup differences in CBF in the present study may also be related to the choice of CBF metrics and the spatial definition of ROI. Previous studies have adopted relative cerebral blood flow (rCBF), calculated as the ratio of the affected hemisphere to the contralateral hemisphere or to a reference region, to reduce interindividual physiological variability in absolute perfusion values, and this approach has been considered helpful in revealing regional hemodynamic abnormalities in certain settings.^20, 21^ In contrast, we analyzed absolute CBF values, which reflect actual perfusion levels but may be influenced by systemic hemodynamic factors such as cardiac output and vascular reactivity, thereby potentially masking subtle intergroup differences.

Moreover, ROIs in the present study were defined according to major arterial territories (MCA, ACA, and PCA). Although this strategy provides clear anatomical correspondence, it may not fully capture perfusion abnormalities in border-zone regions or small focal areas, which may be more susceptible to hemodynamic impairment in chronic ischemic conditions. Therefore, the absence of significant intergroup blood flow differences detected by static ASL in this study does not necessarily imply identical perfusion states between the two groups. The finding of “no significant blood flow differences but directional trends” in this study suggests that for perfusion assessment in chronic occlusive diseases, single-phase ASL results still require optimization through post-processing algorithms and further application of multi-PLD sequences to obtain more comprehensive and accurate perfusion information.

Given that perfusion assessments in chronic occlusive diseases may be influenced by factors such as arterial delivery delays, relying solely on blood flow parameters remains insufficient to fully reflect the true functional state of brain tissue. Therefore, supplementary analysis at the metabolic level is necessary. Metabolic analysis in this study revealed significantly lower or trending lower CMRGlu values in the surgical group, while no difference in SUVR was observed between groups. This phenomenon may arise because CMRGlu reflects the absolute glucose metabolism rate per unit brain tissue, making it more sensitive to metabolic reduction. In contrast, SUVR is a relative metric more susceptible to variations in reference region metabolism, perfusion status, and inter-individual variability.^22–24^ In non-human primate studies, Goutal et al. found that while CMRGlu and SUVR are highly correlated, the former exhibits greater variability and sensitivity to metabolic changes, whereas the latter’s normalization significantly reduces variability.^25^ Laursen et al. observed in severe traumatic brain injury (TBI) patients that quantitative CMRGlu detected global metabolic decline, whereas SUVR normalized to cerebellar metabolism failed to distinguish patients from controls in intergroup comparisons.^26^ These findings suggest that in cases of extensive or bilateral metabolic alterations caused by chronic large vessel occlusion, SUVR may underestimate true metabolic differences, whereas CMRGlu is more suitable as an indicator for evaluating energy compensation status. Further analysis of the supplementary table reveals that patients in the medical therapy group exhibited smaller CMRGlu differences between the ischemic and non-ischemic hemispheres, suggesting relatively balanced bilateral metabolism that may reflect well-preserved collateral circulation and metabolic compensation. In contrast, the surgical group not only demonstrated more pronounced metabolic decline in the ischemic hemisphere but also showed consistently lower CMRGlu values in the non-ischemic hemisphere compared to the medical therapy group, indicating a trend toward widespread metabolic reduction. In patients with chronic intracranial large vessel stenosis or occlusion, collateral circulation may partially maintain relatively stable CBF levels. However, microcirculatory perfusion efficiency, glucose transport, and neuronal metabolic load have already undergone alterations.^27–29^ At this stage, CBF may provide limited reflection of physiological heterogeneity, whereas CMRGlu can more sensitively reveal compensatory exhaustion. This retrospective observational study selected treatment based on real-world clinical judgment rather than PET findings. Although CBF differences between groups were non-significant, patients who ultimately underwent surgery exhibited significantly lower baseline CMRGlu. This suggests that among patients with comparable CBF, the degree of metabolic reduction may more closely align with the “higher-risk” state identified by clinicians. Therefore, when two patients exhibit similar degrees of perfusion decline, CBF alone may be insufficient to determine compensatory capacity. Conversely, a decline in CMRGlu may indicate compromised energy reserves and unsustainable neuronal functional load, placing such patients closer to those clinically considered for revascularization. These findings provide preliminary evidence that metabolic status may hold therapeutic stratification value. Among patients with comparable degrees of blood flow impairment, PET metabolic imaging reveals physiological heterogeneity uncaptured by conventional perfusion assessments. This suggests that future prospective studies should further explore whether baseline CMRGlu could serve as a candidate imaging biomarker for evaluating surgical treatment selection.

However, this study has several limitations that warrant caution when interpreting the results. First, as a single-center retrospective observational study, treatment selection was influenced by multiple factors including symptom severity, hemodynamic assessment, and patient preference. Therefore, it cannot establish a causal relationship between metabolic abnormalities and surgical benefit. Second, the CMRGlu and CBF calculation and normalization methods employed in this study remain relatively simplified. There is room for optimization in utilizing multi-PLD information and post-processing algorithms, which may affect the precision of perfusion and metabolic quantification and the ability to detect subtle differences. Finally, although clinical follow-up was conducted, the primary outcome measure was the mRS functional score. The absence of follow-up perfusion and metabolic imaging data precludes assessment of the dynamic relationship between baseline CMRGlu or CBF and imaging improvement, revascularization efficacy, or metabolic reversibility. It also prevents determination of whether baseline metabolic status truly predicts the extent of postoperative cerebral perfusion and metabolic recovery. These limitations suggest that future studies with larger samples, multiple parameters, prospective designs, and comprehensive follow-up are needed to further validate the potential value of PET metabolic imaging in treatment selection.

## CONCLUSION

This study retrospectively compared baseline perfusion and metabolic status in patients with ICA or MCA steno-occlusive using integrated PET/MRI imaging. Results showed that although no significant difference in CBF levels was observed between the surgical and medical groups, patients in the surgical group exhibited lower CMRGlu, suggesting that metabolic indicators better reflect cerebral tissue energy compensation status than perfusion parameters. Under conditions of similar overall blood flow reduction, decreased metabolism may indicate more limited compensatory capacity. This imaging feature aligns with the patient population clinically selected for revascularization. Findings from this study provide preliminary evidence for the potential value of FDG-PET metabolic imaging in treatment strategy stratification.

## Data Availability

The data that support the findings of this study are available from the corresponding author upon reasonable request.

## Acknowledgments

The authors thank all the patients who participated in this study, as well as the technical staff and nursing team of the department for their valuable support.

## Supplemental Material

Table S1- S7

Figure S1- S2

## Non-standard Abbreviations and Acronyms

ACA: Anterior Cerebral Artery
ASL: Arterial Spin Labeling
CBF: Cerebral Blood Flow
CMRGlu: Cerebral Metabolic Rate of Glucose
EC-IC: Extracranial–Intracranial
FDG: ^18^F-Fluorodeoxyglucose
MCA: Middle cerebral artery
MNI: Montreal Neurological Institute
PCA: Posterior Cerebral Artery
PET/MRI: Positron Emission Tomography and Magnetic Resonance Imaging
PLD: Post-Labeling Delay
ROI: Region of Interest
SUVR: Standardized Uptake Value Ratio

